# Silk fabric as a protective barrier for personal protective equipment and as a functional material for face coverings during the COVID-19 pandemic

**DOI:** 10.1101/2020.06.25.20136424

**Authors:** Adam F. Parlin, Samuel M. Stratton, Theresa M. Culley, Patrick A. Guerra

## Abstract

**Background:** The worldwide shortage of single-use N95 respirators and surgical masks due to the COVID-19 pandemic has forced many health care personnel to prolong the use of their existing equipment as much as possible. In many cases, workers cover respirators with available masks in an attempt to extend their effectiveness against the virus. Due to low mask supplies, many people instead are using face coverings improvised from common fabrics. Our goal was to determine what fabrics would be most effective in both practices.

**Methods and findings:** We examined the hydrophobicity of fabrics (silk, cotton, polyester), as measured by their resistance to the penetration of small and aerosolized water droplets, an important transmission avenue for the virus causing COVID-19. We also examined the breathability of these fabrics and their ability to maintain hydrophobicity despite undergoing repeated cleaning. Tests were done when fabrics were fashioned as an overlaying barrier and also when constructed as do-it-yourself face coverings. As a protective barrier and face covering, silk is more effective at impeding the penetration and absorption of droplets due to its greater hydrophobicity relative to other tested fabrics. Silk face coverings repelled droplets as well as masks, but unlike masks they are hydrophobic and can be readily sterilized for immediate reuse.

**Conclusions:** Silk is an effective hydrophobic barrier to droplets, more breathable than other fabrics that trap humidity, and are readily re-useable via cleaning. Therefore, silk can serve as an effective material for protecting respirators under clinical conditions and as a material for face coverings.

## Introduction

Personal protective equipment (PPE), specifically N95 respirators and surgical masks, are vital to protect against viral transmission during the current COVID-19 pandemic, yet global shortages of these items will likely continue in many locations for the foreseeable future.

Although respirators and masks used by health care providers (HCP) and essential workers (EW) form part of the critical armament against COVID-19, a significant drawback of PPE are that they are purposed for only single use. Sterilization of PPE, especially respirators, has been implemented to enable their continued and repeated use, but this approach reduces the ability of respirators to effectively block particles, can induce damage, or may render the equipment unsafe for further usage [1].

In some cases, HCPs and EWs may only have a single respirator provided to them at their workplace and must reuse them indefinitely under hazardous work conditions. To prolong the life of respirators, many HCPs have adopted the clinical practice of wearing multiple pieces of PPE simultaneously, e.g., a mask on top of a respirator (Fig 1A) [2-4]. This strategy is unsustainable as increased thickness hampers breathing [3] and increases moisture near the wearer’s face, thus becoming a conduit for viral transmission [5,6]. Masks also cannot be adequately cleaned without compromising their protective properties. In many cases, HCPs and EWs remain vulnerable as they have resorted to using (and reusing) less efficient masks on their own when respirators are unavailable, leaving them at greater risk to viral transmission.

**Fig 1.**
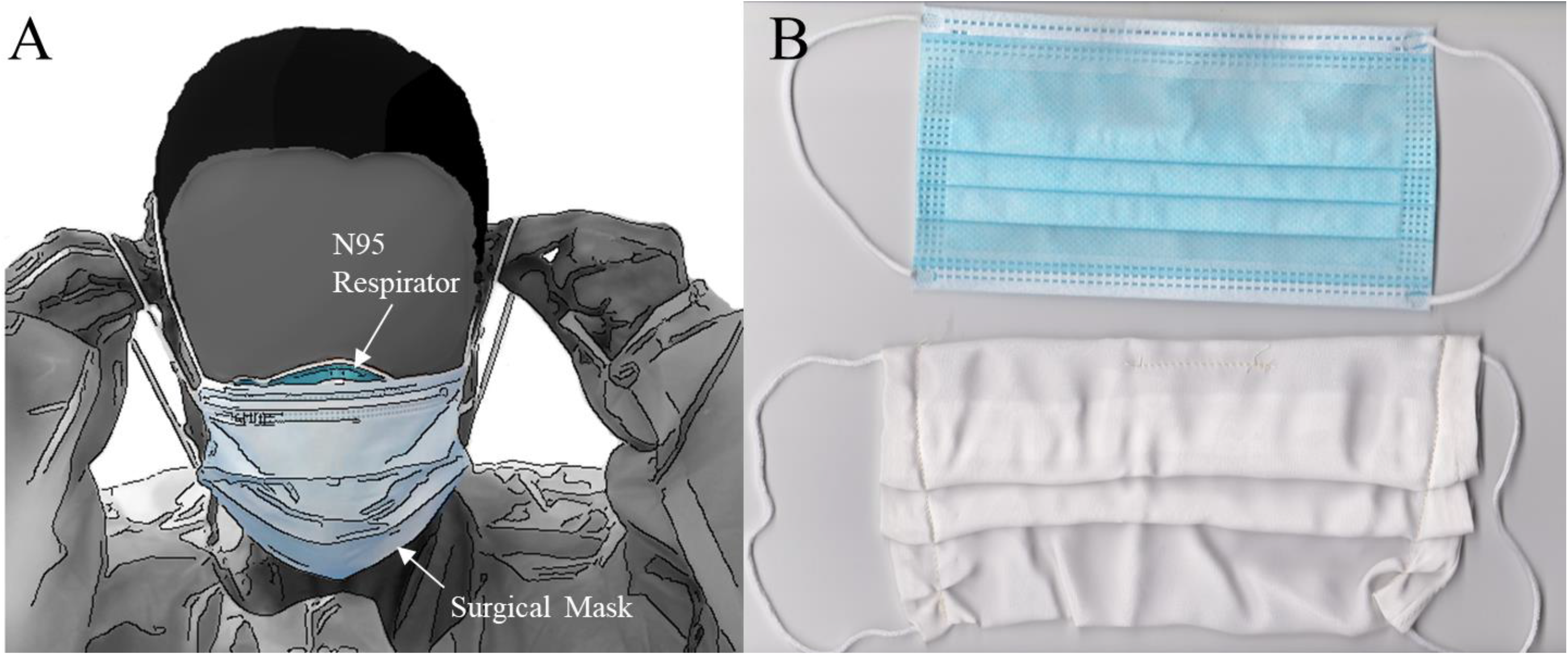
Recommended clinical practice with personal protective equipment (PPE) and a comparison of PPE with a handmade face covering. (A) One commonly used clinical method to prolong and preserve N95 respirators is to layer a surgical mask over it [4]. (B) A commercially available, task-specific surgical mask (top) compared to a silk face covering (bottom) made according to design specifications by the Center for Disease Control and Prevention (CDC) [7]. Photo in (A) was taken by Elaine Thompson (Associated Press) and used with permission.

PPE shortages are now affecting the general population, especially employees instructed to wear masks in the workplace as well as people in public places where mask wearing is mandatory or strongly recommended as part of public health policy [7,8]. As a result, the majority of the general public has been reduced to using improvised face coverings constructed from commercially available materials (Fig 1B) [9,10]. Although the primary purpose of face coverings is to minimize potential viral transmission from the wearer to others [11], they can also provide some protection to the wearer from external sources [12,13]. Cloth, such as cotton, has been suggested as a suitable material for face coverings [14,15], but it remains an open question as to what material possesses the best suite of characteristics to block droplets and viral particles, as well as what material best facilitates comfort, wearability, and reuse of face coverings.

To help combat the PPE shortage amid the COVID-19 pandemic, we examined what materials can serve as immediate solutions for (a) fashioning effective, protective barrier layers that serve to increase the longevity of respirators and the effectiveness of masks under clinical conditions, and (b) for the construction of face coverings to be worn according to current public health guidelines when standard PPE is not available. We focused on the properties of silk, a material commonly used in the garment industry, which is a natural, fibrous biomaterial produced by animals such as moths and spiders. For example, silk moth caterpillars, such as those of the domesticated silk moth, *Bombyx mori*, and of the Robin moth, *Hyalophora cecropia*, produce and use silk for spinning their cocoons [16-18]; these structures consist of hydrophobic and semi-impermeable membranes [19,20] that protect the developing moth residing inside from harsh environmental conditions [20-22]. In addition, protein fibers in silk have been shown to have antimicrobial, antibacterial, and antiviral properties [23-26]. Silk is already used in biomedical applications such as surgical sutures [26], and current research on silk has examined its utility as a biomaterial for many biomedical and human health applications [27-28]. Previous work examining the use of commercially available fabrics for improvised face coverings has also shown that silk possesses some capacity as an antimicrobial barrier when used alone for the fabrication of face coverings [29] and has increased filtration efficiency with more layers [13].

## Methods

### Experimental design

We tested silk material relative to other commercially available fabrics as a protective barrier layer over existing PPE (Fig 1A) and as a material for the construction of face coverings (Fig 1B). We evaluated five types of material that consisted of animal-derived silk that was natural and unmanipulated (i.e., cocoon walls of *B. mori* and *H. cecropia*) or processed (unwashed and washed 100% mulberry silk from pillowcases), processed plant-derived (100% cotton) and synthetic (polyester) fabrics, as well as a water-absorbent material as a positive control (paper towels, see S1 File for material details). These processed fabrics (silk, cotton, and polyester) represent commonly available materials that can be readily used for making protective layers and face coverings. For processed silk, we tested both washed and unwashed silk to examine if the material properties of silk might be altered by standard cleaning techniques (i.e., washing).

We compared the different fabric groups in their level of hydrophobicity, functionally characterized by their ability to block either water or aerosolized droplets (from spray), vehicles for the transmission of the virus underlying COVID-19 [30]. Greater hydrophobicity was defined as the starting contact angles of droplets being greater than 90°, which produces increased resistance to the penetration of droplets into the fabric. We assessed hydrophobicity by first measuring the contact angle behavior of an individual water droplet (5 µL and 2 µL volumes) deposited onto the surface of these materials using the sessile drop technique. In these tests, greater contact angles that are more consistent over time indicate greater hydrophobicity. We also measured the saturation propensity of a droplet (2 µL) and the rate of gas exchange over a 24-hour period through the material to examine the ability of water (liquid and vapor) to penetrate through the material. Gas exchange rates are a measure of porosity and therefore breathability. We also compared the performance of single and multi-layered silk in saturation trials. Finally, we compared the different fabric types and commercially available surgical masks, in terms of penetration of aerosolized droplets delivered as spray through the material, via a modified custom apparatus [31]. We also tested vertical aerosolized spray after sterilization where face coverings were sterilized a total of five times using a dry-heat oven at 70 °C. In all experiments, we tested three different sources for each material type and performed three technical replicates for each source of fabric. Thickness measurements were made in three separate locations on the material and fabric then averaged. More details on these methods, tests, and sterilization are found in S1 File.

### Data analysis

For contact angle trials (both larger 5 µL and smaller 2 µL droplets), we compared the different material types in terms of their starting, dynamic (i.e., change over time), and final contact angles during trials, and the magnitude change in contact angle between the start and final measurements. We analyzed starting and dynamic contact angles, and the magnitude change in contact angle, using a two-way ANCOVA with material thickness as a covariate. Dynamic contact angle data were analyzed using a linear mixed-effect model with *group* and *time* as a fixed effect interaction, and *fabric sample* as the random effect. Individual models were compared against a null using a likelihood ratio test, and the conditional and marginal r^2^ are reported for each model [32]. We analyzed saturation propensity using a two-way ANCOVA with material thickness as a covariate. Gas exchange data were first log_10_-transformed to meet assumptions of normality, and then compared among material types using a one-way ANOVA.

Comparisons of the percentage of samples that were penetrated by a 2 µL water droplet for either single or multilayered silk fabric layers were analyzed using a Fisher’s Exact omnibus test, which was then followed by pairwise Fisher’s Exact tests with Bonferroni correction (α = 0.016). Relative to when no face covering was present over a testing surface, we compared the capability of face coverings (cotton, silk, and polyester) and surgical masks to repel aerosolized droplets (i.e., resist penetration and saturation by aerosol droplets delivered via spray) using a one-way ANOVA. All data were analyzed in R [33]. Significance was set to α = 0.05 except when adjusted for multiple pairwise comparisons.

## Results

### Testing the performance of material for use as protective barriers and face coverings

We found that material groups differed significantly in starting contact angles for both droplet volumes tested (5 µL – ANCOVA: F_6,55_=16.88, P<0.001; η^2^=0.62, η_p_^2^=0.64; 2 µL – ANCOVA: F_6,55_=20.36, P<0.001; η^2^=0.68, η_p_^2^=0.69). In all trials, silk-based materials (*B. mori* and *H. cecropia* cocoons, unwashed and washed silk) were found to be hydrophobic, as they had starting contact angles approximately or greater than 90° (S1 Table). In contrast, cotton, polyester, and paper towel materials were classified as hydrophilic as starting angles were far below 90° or had immediate droplet absorption (S1 Table). Thickness was significantly related to the starting contact angle for both droplet types (5 µL – ANCOVA: F_1,55_=4.47, P=0.039; η^2^=0.03, η_p_^2^=0.08; 2 µL – ANCOVA: F_1,55_=6.87, P<0.05; η^2^=0.04, η_p_^2^=0.11).

In all trials, there was a significant interaction between material group and time for dynamic contact angles (5 µL – *χ*^2^ = 778.58, df=13, P<0.001; marginal r^2^=0.62, conditional r^2^=0.94; 2 µL – *χ*^2^ =549.18, df=13, P<0.001; marginal r^2^=0.46, conditional r^2^=0.93). Hydrophilic materials (cotton, polyester, paper towel), in combination with a lower starting contact angle, had a faster change in contact angle as the droplet was almost immediately absorbed. The droplet stayed on longer, resulting in a gradual change over time, for hydrophobic materials (Fig 2, S2 Table).

**Fig 2.**
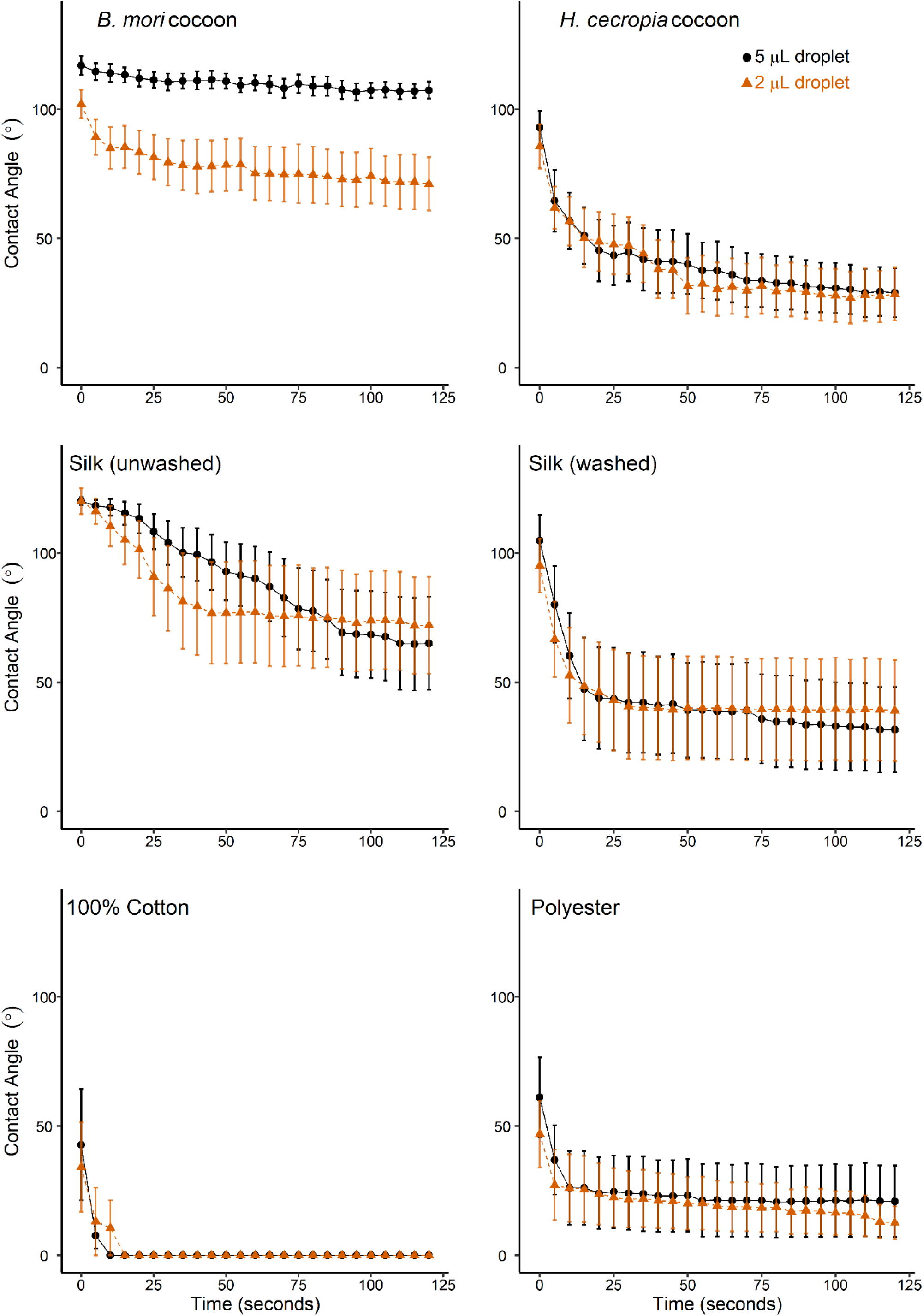
Mean dynamic contact angle (°) of a 5 µL (black) and 2 µL (orange) water droplet for each material group over a 2-minute duration. The positive control (paper towel) is not shown since the water droplet was immediately absorbed and therefore no contact angle could be measured in all trials. For 5 µL droplets, *B. mori, H. cecropia*, washed, and unwashed silk all had starting contact angles above 90° which indicated a hydrophobic surface, while the other fabric types had contact angles less than 90°, indicating a hydrophilic surface. For 2 µL droplets, *B. mori*, washed silk, and unwashed silk all had mean starting contact angles above 90°, which indicates a hydrophobic surface. The remaining fabric groups had contact angles below 90° indicating a hydrophilic surface.

Final contact angles also differed significantly between groups for both droplet volumes tested (5 µL – ANCOVA: F_6,55_=13.02, P<0.001; η^2^=0.62, η_p_^2^=0.64; 2 µL – ANCOVA: F_6,55_=8.72, P<0.001; η^2^=0.52, η_p_^2^=0.56). Thickness was significantly related to the final contact angle for all droplet trials (5 µL – ANCOVA: F_1,55_=25.04, P<0.001; η^2^=0.16, η_p_^2^=0.31; 2 µL – ANCOVA: F_1,55_=19.43, P<0.001; η^2^=0.15, η_p_^2^=0.26; S1 Table).

The magnitude of change from start to final contact angles was significantly different across material groups for all trials (5 µL – ANOVA: F_6,56_=3.48, P<0.01; η^2^=0.27; 2 µL – ANOVA: F_6,56_=3.93, P<0.01; η^2^=0.30). Post hoc pairwise comparisons, however, indicated only significant differences involving paper towel with each other material group (S1 Table).

The saturation propensity of a 2 µL water droplet significantly differed between material groups (ANCOVA: F_6,49_=55.875, P<0.001; η^2^=0.74, η_p_^2^=0.87), with cotton and paper towel having the largest droplet spread followed by the remaining groups (Table 1). Thickness was significantly related to droplet area (ANCOVA: F_1,49_=7.14.884, P<0.001; η^2^=0.03, η_p_^2^=0.23), with a significant interaction between thickness and fabric type (ANCOVA: F_6,49_=9.772, P<0.001; η^2^=0.13, η_p_^2^=0.54). Gas exchange, a proxy for porosity, significantly differed between groups (ANOVA: F_6,56_=16.643, P<0.001, η^2^=0.64). *B. mori* cocoons and cotton material had the highest mean gas exchange rates relative to the other groups (Table 1).

**Table 1.** Saturation (mm^2^) from a 2 µL droplet for material groups where absorption was present (100% cotton, positive control, unwashed silk, synthetic) and not present (*B. mori, H. cecropia*, washed silk) after 60 seconds, and gas exchange rates after a 24-hour period.

| Material Group | Saturation Area (mm<sup>2</sup>) ± SE | Permeability (g/m · s · Pa) ± SE |
| --- | --- | --- |
| *B. mori* cocoon silk layer | 1.59 ± 0.12<sup>c</sup> | 1.92<sup>-09</sup> ± 1.81<sup>-10</sup>, a |
| *H. cecropia* cocoon silk layer | 4.98 ± 1.10<sup>c</sup> | 7.03<sup>-10</sup> ± 7.58<sup>-11</sup> c,d |
| Silk (Unwashed) | 11.96 ± 7.23<sup>b,c</sup> | 8.74<sup>-10</sup> ± 1.03<sup>-11</sup> c,d |
| Silk (Washed) | 5.06 ± 1.47<sup>c</sup> | 9.45<sup>-10</sup> ± 1.45<sup>-11</sup> c,d |
| 100% Cotton | 86.52 ± 17.67<sup>a</sup> | 1.35<sup>-09</sup> ± 6.85<sup>-11</sup> a,b |
| Synthetic | 26.26 ± 10.94<sup>b</sup> | 8.44<sup>-10</sup> ± 1.06<sup>-11</sup> b,c |
| Positive Control (paper towel) | 69.69 ± 22.82<sup>a</sup> | 7.00<sup>-10</sup> ± 2.58<sup>-10</sup> d |

There were significant differences in saturation area between the material groups. The covariate, thickness (mm), was significantly related to the saturation area of each material. For gas exchange rates, silk is as porous as synthetic materials and less porous than 100% cotton. A one-way ANOVA test indicated significant difference between material groups. Groups that share the same letters are not statistically different from each other (Tukey HSD post-hoc tests).

### Single versus multilayered face coverings

To test the resistance of silk layers, we compared the ability of a 2 µL water droplet to penetrate single and multiple fabric layers. We found that droplet penetration of silk fabric significantly decreased as the layers of silk increased from a single layer to either double or triple layers (Fisher’s Exact, P<0.001), but two and three layers of silk did not differ from each other (S3 Table). As the public typically wears face coverings with one or two layers, we compared the capability of single and double layers for silk (washed and unwashed), cotton, and polyester fabrics to resist penetration by aerosolized droplets. Vertical spray tests revealed significant differences between each of the fabric groups and the control of no face covering, when face coverings had one-layer (ANOVA: F_5,42_=18.66, P<0.001; η^2^=0.69) or two-layers (ANOVA: F_5,42_=29.50, P<0.001; η^2^=0.78). There were no differences between any fabric groups, however.

### Exposure of face coverings and masks to vertical aerosolized spray after sterilization

To examine the effects of sterilization, we compared face coverings made from our different test fabrics using recommendations from the CDC [7] with surgical masks. Discoloration of the test surface from the aerosolized spray remained the same for all tested groups with no sterilization (ANOVA: F_4,49_=0.99, p=0.42), one sterilization (F_4,49_=0.98, p=0.43), and five sterilizations (F_4,49_=1.702, p=0.17). This occurred despite significant differences in the thickness of the different face coverings and masks (ANOVA: F_3,41_=713, p<0.001; η^2^=0.98). Cotton face coverings were the thickest (0.367 ± 0.004 mm), followed by masks (0.341 ± 0.008), silk (0.306 ± 0.005 mm) and then polyester (0.216 ± 0.008 mm).

## Discussion

Protective layers and face coverings made from 100% silk, a naturally produced commonly available material, are hydrophobic and can effectively impede the penetration and absorption of both liquid and aerosolized water droplets. The hydrophobicity of silk fabric is further enhanced when used in multiple layers, which when combined, are still thinner than most cotton materials and standard PPE such as surgical masks. Our results demonstrate that the greater hydrophobicity of silk relative to other fabrics, such as cotton and polyester, can make it more effective at impeding droplets, which is a common transmission pathway for the virus that underlies COVID-19 [30].

Silk performs similarly to surgical masks when layered over respirators, as they would occur in clinical settings (Fig 1A), yet has the added advantages of greater hydrophobicity and the ability to be easily cleaned through washing for multiple uses. Recent work has also aimed at making synthetic, reusable hydrophobic layers to layer on top of respirators [34]. The use of natural silk barriers to protect PPE adds to these initiatives, but with the added benefits of silk’s inherent beneficial properties and accessibility of silk for both commercial and public use. Here, the sericulture, textile, and garment industries, along with their supply networks and infrastructure, potentially have a direct pathway to becoming important partners against the current COVID-19 pandemic and in future public health emergencies in which PPE may again be in short reserve.

A limitation of respirators and masks, but especially face coverings, is that normal breathing can be hampered when worn, and this difficulty increases with thickness. Prolonged use also exposes individuals to added risks, as they increase the local humidity around the area upon which it is worn (>90% relative humidity) [35], thereby creating a potential pathway for the virus to travel due to the trapped moisture near the face that inadvertently increases wetness [5,36]. Increased humidity underneath these items, exacerbated when worn in hot and humid environments, significantly decreases their wearability because of higher friction and skin moisture [37]. This creates discomfort and can result in an individual unintentionally touching their face. Our results suggest that this limitation and its accompanying risks can be mitigated by silk, at least when it is used in the fabrication of face coverings.

Currently, public health recommendations focus on cotton material for face coverings [14]. We found that cotton materials are hydrophilic, and readily allow droplets to rapidly penetrate and saturate the fabric like a sponge. Therefore, face coverings made out of these materials may quickly become reservoirs of virus and act as conduits for viral transmission when worn, even after a short time [5,6]. Face coverings made out of polyester face these same limitations, as it is hydrophilic like cotton. Therefore, cloth and polyester face coverings appear to be more suitable for brief, one-time use. In contrast, silk’s hydrophobicity, increased filtering efficiency when layered [13], and lack of capillary action [20] make it a more advantageous material for face coverings that are also thin, light, and breathable. Recent recommendations by the World Health Organization have also mentioned combining hydrophilic and hydrophobic layers when creating face coverings [38], and our work supports the use of silk as a better hydrophobic layer for face coverings that is more effective than either cotton or polyester material that are hydrophilic. Furthermore, our results also suggest that using multiple layers of silk for face coverings can increase filtering efficiency, yet retain and enhance its advantageous hydrophobic properties that can preclude it from becoming a reservoir and conduit for the virus, while remaining breathable and comfortable when worn.

Although N95 respirators are still the most effective form of protection against viral transmission, our study highlights the practicality of using current commercially available 100% silk material as a protective barrier for PPE and as a material for face coverings. Moreover, silk may play a major role in the development of new PPE equipment, such as respirator inserts, that capitalize on its many benefits. For example, silk possesses antimicrobial, antiviral, and antibacterial properties [24,26], potentially due to the presence of copper, a compound that has antiviral properties and which animals naturally incorporate into their silk [23]. Other fabrics and non-specialized PPE require copper particles to be infused during the manufacturing process [39], an expensive process that could be circumvented by using natural silk fibers. In summary, we suggest that silk has untapped potential for use during the current shortage of PPE in the ongoing COVID-19 pandemic. It can be effective when used as a covering to extend the lifetime of N95 respirators, when fashioned as face coverings for the general public, and as a material for the development of the next generation of PPE.

## Data Availability

All relevant data are found within the paper and its Supporting information files.

## Abbreviations

EW: essential worker
HCP: health care provider
PPE: personal protective equipment

## Acknowledgements

We thank Eric J. Tepe for logistical support and helpful comments during the course of this study. P.A.G., A.F.P., T.M.C. and S.M.S. were supported by funds from the University of Cincinnati.

## Data reporting

All relevant data are found within the paper and its Supporting information files.

## Supporting information

**S1 Fig.**
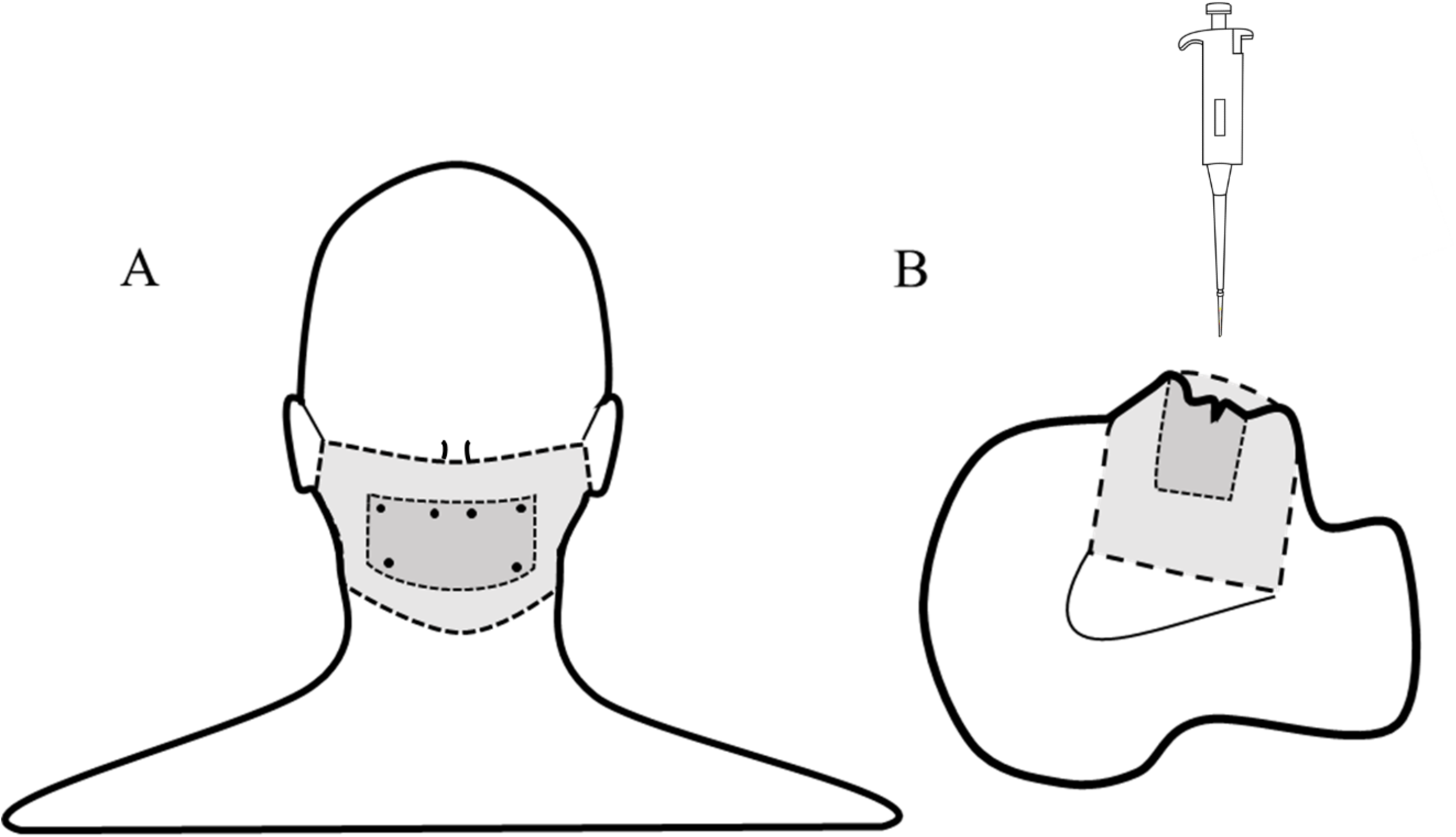
Experimental set-up and notecard attachment for the sessile horizontal droplet tests. Material barriers were held to the mannequin head using pins. The dark-grey covering represents the note-card placement while the light-grey represents the face covering.

**S2 Fig.**
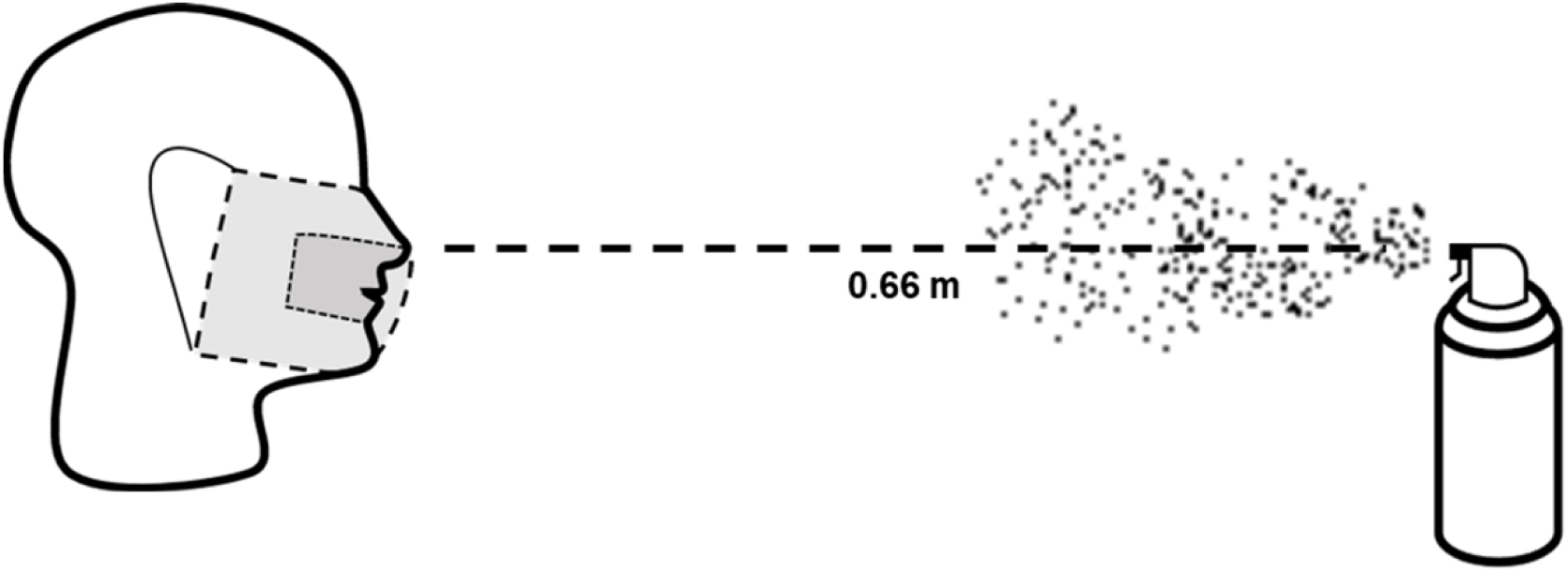
Aerosolized spray experimental set-up with mannequin head (no mask or face covering during trials) and aerosolizing apparatus. Prior to each test, the apparatus was filled to 82 kPa. The dark-grey covering represents the note-card placement while the light-grey represents the face covering.

**S1 Table.**
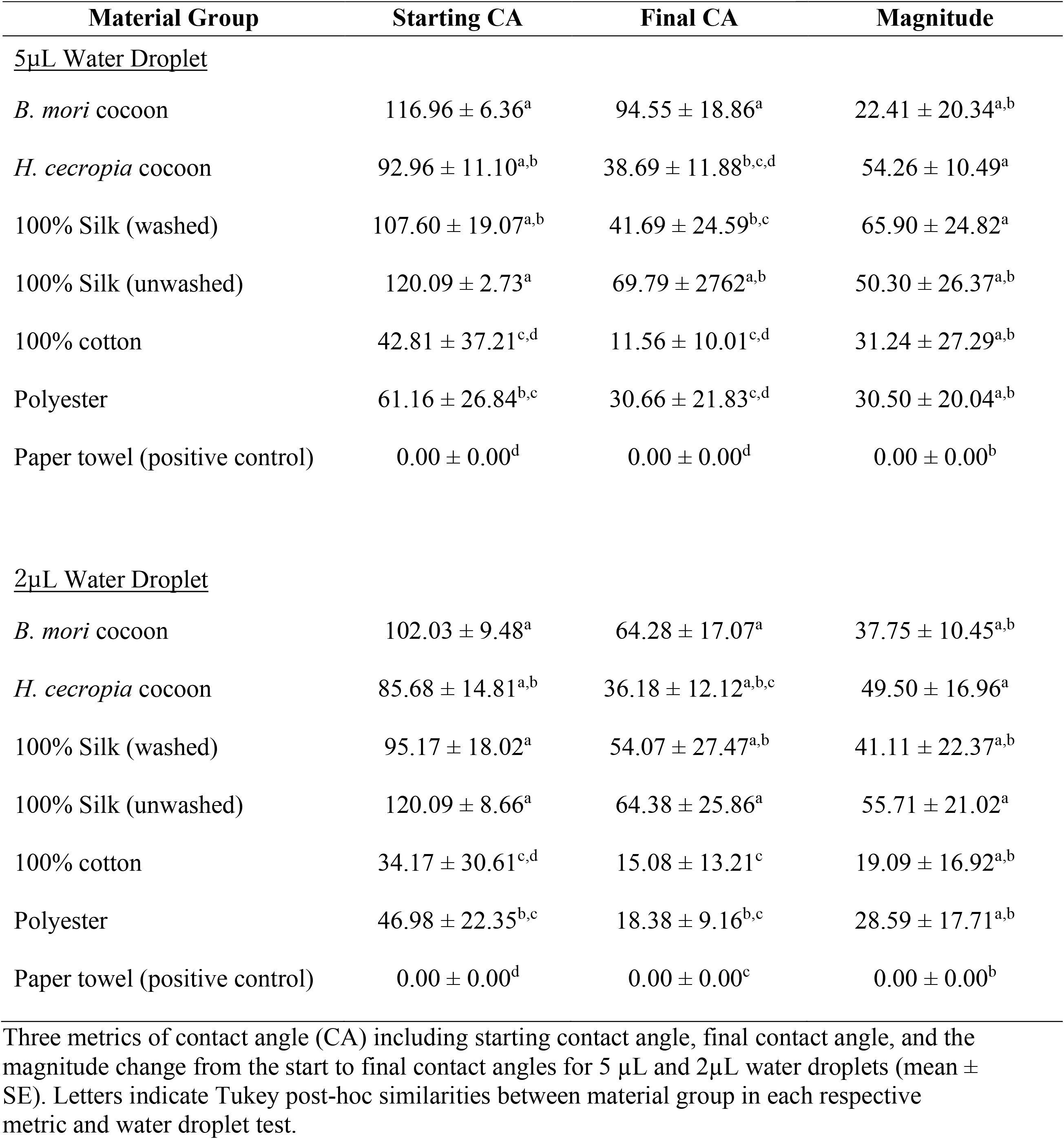
Contact angle metrics. Three metrics of contact angle (CA) including starting contact angle, final contact angle, and the magnitude change from the start to final contact angles for 5 µL and 2µL water droplets (mean ± SE). Letters indicate Tukey post-hoc similarities between material group in each respective metric and water droplet test.

**S2 Table.** Summary of mixed-effect models for dynamic contact angle of a 2 µL and 5 µL water droplets.

| *Predictors* | **2μL Contact Angle** |  |  | **5μL Contact Angle** |  |  |
| --- | --- | --- | --- | --- | --- | --- |
|  | *Estimates* | *CI* | *P* | *Estimates* | *CI* | *P* |
| (Intercept) | 87.72 | 66.49 – 108.96 | <0.001 | 114.13 | 95.10 – 133.16 | <0.001 |
| Time | -0.16 | -0.20 – -0.12 | <0.001 | -0.07 | -0.11 – -0.02 | 0.004 |
| *H. cecropia* | -29.88 | -59.91 – 0.15 | 0.051 | -54.74 | -81.66 – -27.82 | <0.001 |
| 100% Cotton | -79.31 | -109.34 – -49.28 | <0.001 | -106.58 | -133.50 – -79.66 | <0.001 |
| Paper Towel | -87.72 | -117.76 – -57.69 | <0.001 | -114.13 | -141.05 – -87.21 | <0.001 |
| 100% Silk (unwashed) | 16.02 | -14.01 – 46.06 | 0.296 | 6.24 | -20.68 – 33.16 | 0.649 |
| 100% Silk (washed) | -31.64 | -61.67 – -1.61 | 0.039 | -51.63 | -78.55 – -24.71 | <0.001 |
| Polyester (synthetic) | -58.49 | -88.52 – -28.45 | <0.001 | -81.81 | -108.73 – -54.89 | <0.001 |
| Time * *H. cecropia* | -0.16 | -0.23 – -0.10 | <0.001 | -0.24 | -0.31 – -0.18 | <0.001 |
| Time * 100% Cotton | 0.06 | -0.00 – 0.12 | 0.069 | -0.03 | -0.09 – 0.04 | 0.443 |
| Time * Paper Towel | 0.16 | 0.10 – 0.22 | <0.001 | 0.07 | 0.00 – 0.13 | 0.043 |
| Time * 100% Silk (unwashed) | -0.18 | -0.24 – -0.11 | <0.001 | -0.45 | -0.51 – -0.38 | <0.001 |
| Time * 100% Silk (washed) | -0.04 | -0.10 – 0.03 | 0.248 | -0.26 | -0.32 – -0.19 | <0.001 |
| Time * Polyester | 0.02 | -0.05 – 0.08 | 0.591 | -0.06 | -0.13 – 0.00 | 0.052 |
| Marginal R<sup>2</sup> / Conditional R<sup>2</sup> | 0.463 / 0.932 |  |  | 0.619 / 0.938 |  |  |

**S3 Table.** Percentage of 2 µL water droplets that penetrated single, double, or triple silk layers compared with Fisher’s Exact test. Totals (n=6) are from both washed (n=3) and unwashed (n=3) silk materials where four 2 µL droplets were applied per technical replicate. Identical letters indicate similarities between Bonferroni-corrected pairwise comparisons.

| Thickness | Penetrated | Total Droplets | Percent (%) |
| --- | --- | --- | --- |
| 1-Layer | 34 | 72 | 47.2<sup>a</sup> |
| 2-Layer | 2 | 72 | 2.78<sup>b</sup> |
| 3-Layer | 1 | 72 | 1.39<sup>b</sup> |

### S1 File. Protocol for experiments

#### Materials and fabrics tested

We tested five material groups for contact angle, saturation propensity, and gas exchange rates, then tested three fabric groups for droplet penetration of single and multiple layers, and for resistance to aerosolized spray. For animal-derived silk that was natural and unmanipulated, we took *Bombyx mori* cocoon samples from our current laboratory colony (3^rd^ generation reared; Department of Biological Sciences, University of Cincinnati) and *Hyalophora cecropia* cocoon samples collected outdoors from Eastern and Central, Massachusetts between 2013-2016 [1]. For animal-derived processed silk materials, we tested black (unwashed thickness: 0.094 ± 0.002 mm; washed: 0.092 ± 0.003 mm) and white (unwashed thickness: 0.0.112 ± 0.001 mm; washed: 0.103 ± 0.004 mm) 100% silk scarves (Cyzlann, Shenzhen Yirong Technology Co. Ltd., Shenzhen, Guangdong, China) and a 100% mulberry silk (unwashed thickness: 0.165 ± 0.001 mm; washed: 0.169 ± 0.003 mm) pillowcase (Ravmix, Shenzhen City, Yuanmi Trade Co., Ltd., Guangdong, China). Subsets of the silk material were washed according to instructions outlined by the distributor to create the silk (washed) group. For processed plant-derived material (100%), we tested a 100% cotton handkerchief (thickness: 0.158 ± 0.010 mm), 100% cotton fabric (thickness: 0.199 ± 0.006 mm), and a 100% Egyptian cotton pillowcase (thickness: 0.163 ± 0.005mm). Synthetic materials tested included a pillowcase that was a blend of 88% polyester – 12% nylon (thickness: 0.152 ± 0.002 mm), a 100% polyester pillowcase (thickness: 0.103 ± 0.001 mm), and a 100% polyester drawstring bag (thickness: 0.088 ± 0.005 mm). The 100% Egyptian cotton pillowcase, 100% cotton handkerchief, 100% cotton fabric, 100% polyester pillowcase, 88% polyester-12% nylon pillowcase, and 100% polyester drawstring bag were purchased two-years prior from various retailers (Walmart, USA; JoAnn Fabrics, USA). Positive controls (i.e., paper towels) consisted of a generic brand for white paper towel (thickness: 0.129 ± 0.008 mm; Proctor & Gamble, USA), brown paper towel (thickness: 0.083 ± 0.004 mm; Home Depot, USA), and a Kimwipe (thickness: 0.073 ± 0.004mm; KimTech, USA). Fabrics used for face coverings tested in aerosolized spray experiments were made from 100% mulberry silk material, 100% cotton material, and 100% polyester material. Surgical masks were purchased from local retail stores (surgical mask type 1: Huizhou Canice Health Material Co. Ltd, Guangdong Province, China; surgical mask type 2: Jiangsu Nanfang Medical Co. Ltd, Changzhou, Jiangsu, China).

#### Water droplet contact angle

Contact angle data for 5 µL and 2 µL water droplet trials were collected using an experimental setup based on those used in previous work [2]. The droplet volumes were based on the range of values previously used to test natural materials and fabrics [3,4]. The contact angle of a water droplet deposited onto the material surface (using the sessile drop technique; see below) was used to determine the hydrophobicity of the test material based on the angle produced by the edge of a water droplet contacting the material surface. We vertically deposited the water droplet (5 or 2 µL) onto the fabric piece using a pipette. We avoided any effects of kinetic energy on the contact angle formed by the droplet by ensuring the droplet was in contact with both the pipet tip and the surface of the material swatch prior to final deposition [5]. We used a high-resolution digital camera (Micro 4/3 Lumix SLR, Panasonic Corporation) to capture trial images. During all trials, the camera was kept level with the water droplet and test material. We performed trials on a plastic platform that was positioned horizontally and leveled using a leveler (Bullseye Surface Level, Empire Level). For each trial, we obtained three mean contact angle measurements (mean angle of the contact angle of the left and right sides of the droplet as seen in images): the initial contact angle (time = 0 s, the first image that the pipette tip was completely out of frame), the dynamic contact angle (mean contact angle, sampled every five seconds, and averaged at the end of the trial), and the final (defined as the last reliable image in which the contact angle could be determined or at *t* = 120s). We tested the contact angle of a 5 µL and 2 µL water droplets separately.

Images were sampled every second for a total duration of two minutes and then uploaded to ImageJ 1.52a (http://rsb.info.nih.gov/ij/) for analysis. The two points of contact were then identified as the outer most points at which the droplet touched the material surface. A straight line was then drawn with the angle tool connecting the two points of contact, parallel to the plane of the material, and the angle line was drawn tangential to the point of contact between the droplet and the material. This technique was done for both the right and left side of the droplet and then averaged to obtain the mean contact angle [6]. A contact angle measurement was determined unreliable if either of the two points of contact or the curvature of the droplet could not be determined.

#### Saturation propensity and gas exchange

Saturation propensity was measured as the absorption of a water droplet and was used to test the permeability of the test material. For each trial, we applied a 2 µL water droplet and waited 1-minute before taking an image of the material to measure the total area that the water droplet had spread within the material. Images were analyzed using ImageJ. If the water droplet was not absorbed at the end of 1-minute, we measured the area of the water droplet. The water droplet was applied using a similar technique as above by ensuring the droplet was in contact with the material first before depositing the droplet.

We tested the rate of gas exchange for each material by using methods that were modified from previous studies [7,8]. First, we built an airtight holder for material swatches through which only water vapor was allowed to evaporate. The apparatus was created from a 0.3 mL micro reaction vessel with a hole in the rubber seal to keep the vessel airtight. Each micro reaction vessel was filled with water (300 µL), covered with the material swatch and airtight cap, and then placed on an electronic balance in the room to obtain the initial weight and to measure the weight change after a 24-hour period. We recorded the ambient temperature and humidity of the room for the duration of these tests to correct for the water vapor transfer rate [9].

#### Single and multi-layer silk water droplet absorption

We determined how increasing the number of layers of silk affects the ability of silk to be an effective barrier by comparing the ability of a 2 µL water droplet to penetrate one, two, and three layers of silk fabric for washed and unwashed silk groups. For each trial, we placed a 7.62 cm by 12.70 cm notecard on a Styrofoam mannequin head (Fig. S1a), which covered the nose, mouth, and upper cheek areas (left and right) while the mannequin head was in a horizontal position (Fig. S1b). Each trial was completed when the 2 µL droplet was no longer present on the surface of the silk, either through absorption or evaporation. Images were scanned using a flatbed scanner (Canon MG2220, Canon, Inc.), then uploaded and processed in ImageJ v1.52a. Blank index cards (Walmart Inc., AR, USA) were used to identify possible potential discoloration in the card from the manufacturing process that would create a false positive detection during image analysis. These were identified as small dark points on the card that differed from discoloration caused by the droplet.

#### Standardizing aerosolized spray trials

The velocity of the spray was determined through the relationship of flow rate and velocity using the following equations for flow rate (m^3^/s):

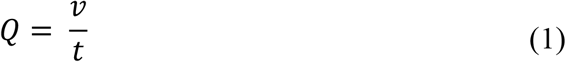

where *Q* is the flow rate (m^3^/s), *v* is the volume (m^3^), and *t* is time (s). The relationship between flow rate and velocity is as follows:

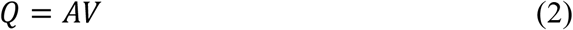

where *Q* is flow rate (m^3^/s), *A* is the cross-sectional area of the cylinder (m^2^), and *V* is the velocity (m/s). We solved for velocity by first calculating the flow rate (*Q*) from equation (1) and then rearranging equation (2). We recorded each spray using a camera (Logitech HD Pro C920) and weighed the apparatus before and after each spray. The aerosolized spray had an average velocity of 0.88 ± 0.04 m/s with each spray containing 0.125 ± 0.05 mL of liquid.

Although a real human cough has an extreme amount of variability in droplet size, cough plume, and other characteristics [10], our device based on a similar experimental design [11] represents an extreme case in which a patient openly coughs in close proximity without any protective barrier.

#### Single and multilayer barrier aerosolized spray test

We compared 100% cotton, polyester, and 100% silk (washed and unwashed) as a single layer and double layer in the aerosolized spray test. We modified an aerosol can with a standard valve, and added 150 ml of black-dyed water (10ml black dye, 140ml water; McCormick, MD, USA). Prior to each trial, the aerosol can was filled to 82 kPa with an air pump and checked using a tire-pressure gauge. The Styrofoam mannequin head had a piece of fabric on top of a notecard pinned to the face (Fig. S1a), and which was placed 0.66m [10] from the aerosol can (Fig. S2). A control group (no mask) was sprayed to provide a baseline of discoloration for comparison. The aerosolized droplets were of a random distribution in size with the speed and total volume consistent across trials.

#### Face covering and mask aerosolized spray test

Face coverings were made according to the CDC guidelines for sewn pleated face coverings [12], and were made with a single material that consisted of either polyester, cotton, or silk (see above). We made three face coverings for each material group and included two brands of surgical masks for comparison in the aerosolized spray test. Initially, these coverings were tested prior to any sterilization and stretching. After the initial trials, the coverings were sterilized using dry heat (70 °C) for 1-hour each and then retested after a single sterilization and after five sterilizations. After each sterilization, face coverings were worn for approximately 5-minutes and stretched (i.e., diagonally, horizontally, and vertically) to simulate wear-and-tear, and the same masks were used across all trials.

Images were processed in ImageJ 1.52a. The images were converted into 16-bit images to allow grayscale thresholding to isolate and separate pixels darkened by the aerosolizing apparatus. Using a positive control, the threshold value was determined by incrementally increasing the value until both visible spots were sufficiently covered and before there was significant threshold identification on either the white of the card or on the background on which the cards were placed. From this process, we were able to obtain an area and associated identity for every contiguous threshold particle. This tool enabled us to exclude any particles that were obviously not droplets from the aerosolizing apparatus and instead resulted from the experimentation itself. These included (1) holes created by the pins securing the card to the fabric, (2) large shadowed portions of the card created by unintentional bending or creasing of the card during experimentation, and (3) large fabric remnants or other debris found on the card. After these areas were excluded, the total sum area of all the threshold particles was calculated.

